# Extracellular matrix stiffness in oral squamous cell carcinoma: a systematic review and meta-analysis of associations with tumor progression and prognosis

**DOI:** 10.64898/2026.09.28.26363990

**Authors:** Igor Felipe Pereira Lima, Ana Carolina Toebe Silva, Adam Jeffrey Engler, Leonardo da Silva Bittencourt, Marcelo Lazzaron Lamers

**Affiliations:** Department of Oral Pathology, School of Dentistry, Federal University of Rio Grande do Sul, Porto Alegre, RS, Brazil; Department of Stomatology, School of Dentistry, Federal University of Rio Grande do Sul, Porto Alegre, RS, Brazil; Chien-Lay Department of Bioengineering, UC San Diego, San Diego, California, USA; Department of Morphological Sciences, Institute of Basic Health Sciences, Federal University of Rio Grande do Sul, Porto Alegre, RS, Brazil; State Technical School of Health, Hospital de Clínicas de Porto Alegre (HCPA), Porto Alegre, Brazil

**Keywords:** Oral squamous cell carcinoma, Extracellular matrix, Tissue stiffness, Biomechanics, Prognosis, Elastography

## Abstract

**Objective:** To critically evaluate the influence of extracellular matrix stiffness on tumor progression, prognosis, and clinical implications in patients with oral squamous cell carcinoma.

**Design:** This systematic review followed the PRISMA (Preferred Reporting Items for Systematic Reviews and Meta-Analyses) guidelines, and the protocol was registered in PROSPERO (CRD420251156429). Five databases (PubMed, Scopus, Embase, Web of Science, and OATD) were searched without restrictions on publication year or language. Human studies that evaluated extracellular matrix stiffness in oral squamous cell carcinoma, either as an assessment method or as a prognostic factor, were included; in vitro and animal studies were excluded.

**Results:** Eight human studies involving 436 patients were included. Elastography was the most frequent measurement method, employed in four studies. Mean tumor thickness was reported in three studies and ranged from 3.0 to 4.4 mm. Extracellular matrix stiffness was expressed in various units (e.g., 37.7 gf, 23.5 kPa) and was consistently higher in tumor tissue than in normal mucosa in all studies. Three studies demonstrated a significant positive correlation between stiffness and tumor thickness. One study identified predictive cut-off values for lymph node metastasis and reported that high matrix stiffness was an independent predictor of worse overall survival (HR = 4.022; 95% CI: 1.294–12.495; p = 0.016) and recurrence-free survival (HR = 5.680; 95% CI: 1.146–28.143; p = 0.033). A meta-analysis of five studies (307 patients) showed a significantly higher standardized stiffness in oral squamous cell carcinoma compared with normal oral mucosa (standardized mean difference = 5.28; 95% CI: 2.61–7.94; p < 0.001), with substantial heterogeneity (I² = 98.6%).

**Conclusions:** Extracellular matrix stiffness is elevated in oral squamous cell carcinoma and is associated with greater tumor thickness, lymph node metastasis and, in one study, worse survival. These findings support extracellular matrix stiffness as a potential biomechanical marker, pending further validation in standardized prospective studies.

## INTRODUCTION

Oral Squamous Cell Carcinoma (OSCC) is one of the most prevalent malignancies of the head and neck and is considered a major global public health concern [1–3]. In 2020, approximately 378,000 new cases were reported worldwide according to Global Cancer Observatory (GLOBOCAN) data, with projections indicating approximately 40% increase in incidence by 2040 [4]. Early detection is critical for improving prognosis and reducing morbidity and mortality [5–7]. However, over 60% of OSCC cases are diagnosed at advanced stages [8], when the 5-year survival rate is reduced [4,8]. This delayed diagnosis largely stems from the subtle nature of early clinical presentation.

A clinically relevant hallmark of malignant transformation in solid tumors is increased tissue stiffness [9]. This mechanical alteration is intrinsically associated with extracellular matrix (ECM) remodeling, whereby the tumor microenvironment becomes significantly stiffer than adjacent normal tissues, primarily due to increased cellular density, intense fibrosis, and qualitative and quantitative changes in ECM composition [9–11]. In some tumor types, this difference can even be detected by manual palpation of the tissue itself. Histologically, ECM stiffness is driven by excessive collagen deposition and crosslinking mediated by enzymes such as lysyl oxidase (LOX), a phenomenon known as desmoplasia [11,12]. Increased stiffness rather acts as an active modulator of tumor cell behavior through mechanotransduction [13,14]. In this process, mechanical signals induced by ECM stiffness are converted into intracellular biochemical responses, activating pathways such as Focal Adhesion Kinase (FAK) and promoting invasion, cell migration, and epithelial-mesenchymal transition (EMT) [12–14].

For objective and non-invasive assessment of these mechanical properties of tumor stiffness, elastography methods have been employed, providing quantitative data on tissue elasticity [12,15]. The clinical utility of these methods is well established in medicine, particularly in breast, thyroid, and hepatobiliary oncology, having demonstrated high diagnostic potential in differentiating between malignant and benign lesions [16–18]. This success is based on the ability of these techniques to quantitatively map the elevated stiffness characteristic of malignant tissues [15,16].

Despite the growing recognition of stiffness as a tumor biomarker and the promising results of assessment methods in other cancer types, its systematic application in the context of OSCC remains incipient [19,20]. In those other cancer types, ECM stiffness is linked to mechanisms of tumor progression, such as the induction of an invasive-dormant subpopulation in response to mechanical stress [20,21], suggesting a crucial role for these methods not only in diagnosis but also in predicting lymph node metastasis [22]. Considering that stiffness measurement can aid in early detection, provide prognostic information, and guide therapeutic decisions [23,24], it is essential to systematically consolidate the available evidence for OSCC. In this context, the present systematic review aims to critically evaluate the influence of ECM stiffness on tumor progression, prognosis, and clinical implications in patients with OSCC.

## MATERIALS AND METHODS

### Protocol and registration

This systematic review was conducted following the Preferred Reporting Items for Systematic Reviews and Meta-Analyses (PRISMA) [25] statement and Cochrane guidelines [26]. The review protocol was registered in the PROSPERO database CRD420251156429.

### Study design

This is a systematic review designed to answer the following guiding PEO question: “Does ECM stiffness (***Exposure***) influence the progression and prognosis (***Outcomes***) of patients with oral squamous cell carcinoma (***Population***)?”

### Eligibility criteria

Human studies evaluating ECM or tissue stiffness in OSCC, either as a diagnostic assessment method or as a prognostic factor, were eligible for inclusion. There were no restrictions on publication year, language, or publication status (including articles in press). The following studies were excluded: (I) studies not assessing ECM or tissue stiffness in OSCC; (II) studies focused on other cancer types without specific, extractable data for oral cancer; (III) review articles, editorials, letters to the editor, and conference abstracts without complete, extractable data; (IV) studies evaluating only ECM composition without any assessment of stiffness; (V) studies in which oral cancer data could not be isolated from other head and neck cancer sites; (VI) animal and in vitro studies.

### Sources of information and search

Two reviewers (IFPL and ACTS) independently searched the electronic databases up to September 2025. Search terms were identified using DeCS (Health Sciences Descriptors), MeSH (Medical Subject Headings), and Emtree (Embase Subject Headings), and combined using the Boolean operators “AND” and “OR” to optimize the search strategy (Table 1). Retrieved records were exported to EndNote Basic/Online (Thomson Reuters, Toronto, Canada), and duplicates were removed prior to study selection.

**Table 1.** Search strategies in the databases.

| *Databases* | *Search strategies (September, 2025)* | *Results* |
| --- | --- | --- |
| *PubMed*<br>*http://www.ncbi.nlm.nih.gov/pubmed* | ("stiffness" OR "extracellular matrix" OR "matrix stiffness") AND ("oral cancer" OR "head and neck cancer") | 397 |
| *Scopus*<br>*https://www.scopus.com/* | ("stiffness" OR "extracellular matrix" OR "matrix stiffness") AND ("oral cancer" OR "head and neck cancer") | 847 |
| *Embase*<br>*https://www.embase.com/* | ('stiffness'/exp OR 'stiffness' OR 'extracellular matrix'/exp OR 'extracellular matrix' OR 'matrix stiffness'/exp OR 'matrix stiffness') AND ('oral cancer'/exp OR 'oral cancer' OR 'head and neck cancer'/exp OR 'head and neck cancer') | 3,735 |
| *Web Of Science*<br>*http://apps.webofknowledge.com/* | ("stiffness" OR "extracellular matrix" OR "matrix stiffness") AND ("oral cancer" OR "head and neck cancer") | 466 |
| *OATD*<br>*https://oatd.org/* | ("stiffness" OR "extracellular matrix" OR "matrix stiffness") AND ("oral cancer" OR "head and neck cancer") | 35 |
| ***TOTAL*** |  | 5,480 |

### Study selection

The selection of studies was conducted in three distinct stages. In the first stage, two reviewers (IFPL and ACTS) systematically screened the titles of all retrieved records. Articles with titles relevant to the objective of this systematic review were selected for the second stage. In the second stage, the same reviewers screened the abstracts and applied the inclusion and exclusion criteria. Studies without available abstracts were evaluated in the third stage. In this final stage, the full texts of the articles were assessed. When disagreements arose regarding a study’s eligibility, a third reviewer (MLL) was consulted to make a final decision. Studies that did not meet the criteria were recorded separately, with the reasons for exclusion clearly documented.

### Process of data collection and extraction

Two authors (IFPL and ACTS) performed data extraction using specific spreadsheets designed for this purpose. The following information was extracted from the eligible studies: author and year, sample size (n), sex, age (years), tumor location, tumor type, tumor stage, follow-up, stiffness measurement method, stiffness classification, tumor thickness (mean), quantitative stiffness parameters (mean ± SD), histopathological correlations, progression factors, and prognostic outcomes. As a calibration exercise, the reviewers (IFPL and ACTS) discussed the eligibility criteria and applied them to a random sample of 20% of the retrieved studies to assess inter-rater agreement. After reaching an adequate level of agreement (κ = 0.83–0.87), the reviewers independently extracted data from all studies. Any discrepancies were resolved by consensus and supervision of a third reviewer (MLL). Authors of studies with insufficient data or missing information necessary for summarization and comparison were contacted via email to request additional details.

### Risk of bias assessment

The risk of bias for individual studies was independently assessed by two authors (IFPL and ACTS) using the “*Joanna Briggs Institute Critical Appraisal Checklist for Diagnostic Test Accuracy Studies*” and “*The Joanna Briggs Institute Critical Appraisal Checklist for Cohort Studies*”. To facilitate summarization, studies were classified as having high, moderate, or low risk of bias based on the proportion of “yes” responses: **high risk** for ≤49%, **moderate risk** for 50–69%, and **low risk** for ≥70% of “yes” responses (Table 3). These thresholds were defined a priori and should be interpreted as an approximate guide, given the lack of validated cut-offs for these checklists (Table 3).

### Data synthesis and statistical analysis

Data from the five of eight included studies were integrated and analyzed using Comprehensive Meta-Analysis (CMA) software, Version 4 (Biostat, Inc., Englewood, NJ, USA; available at meta-analysis.com). The primary outcome measure consisted of continuous data, specifically assessing the standardized mean difference in ECM stiffness between oral control (normal oral mucosa) and OSCC samples. Effect sizes were calculated and presented as standardized mean differences (Cohen’s d) with their respective 95% confidence intervals (CI). Forest plots were generated to visually represent the standardized mean differences and 95% CI for each individual study, along with the pooled summary effect estimate. A distribution plot of true effects was also constructed to illustrate the overall pattern of effect sizes across studies.

Statistical heterogeneity was assessed using the I² statistic, with values of 0–40%, 30–60%, 50–90%, and 75–100% interpreted as negligible, moderate, substantial, and considerable heterogeneity, respectively. In view of the anticipated clinical and methodological diversity (including different stiffness measurement techniques such as elastography, manual devices, and histological quantification), a random-effects model (DerSimonian–Laird method) was used for all meta-analyses. Given the heterogeneity in measurement methods and scales, the pooled SMD should be interpreted as an exploratory summary of the direction and magnitude of differences rather than as a precise estimate of a common effect.

Publication bias was assessed through both visual and quantitative methods. Funnel plots and precision plots were generated to visually identify potential publication bias by plotting the precision of each study against it corresponding standardized mean difference. Asymmetry in the funnel plot, particularly the absence of small studies exhibiting negative effects, may suggest the presence of publication bias. Additionally, Egger’s regression intercept test and Begg’s rank correlation test were employed to statistically assess funnel plot asymmetry, contingent upon the number of studies included and deemed appropriate for such analysis.

## RESULTS

### Selection of studies

Searches of PubMed, Web of Science, Embase, Scopus, and OATD databases resulted in 5,480 articles. After removing duplicates, 3,995 records remained for title screening. Of these, 180 were considered potentially eligible for abstract review. After abstract screening, 23 articles were selected for full-text assessment. In addition, all references of the selected manuscripts (a total of 634 references) were carefully analyzed to identify articles not captured by the main search strategy, resulting in the inclusion of two additional eligible studies. Thus, 25 articles were assessed in full text. After excluding 17 articles for not assessing stiffness (n=10) or for being in vitro studies (n=7), 8 studies were ultimately included in this systematic review (Figure 1).

**Figure 1.**
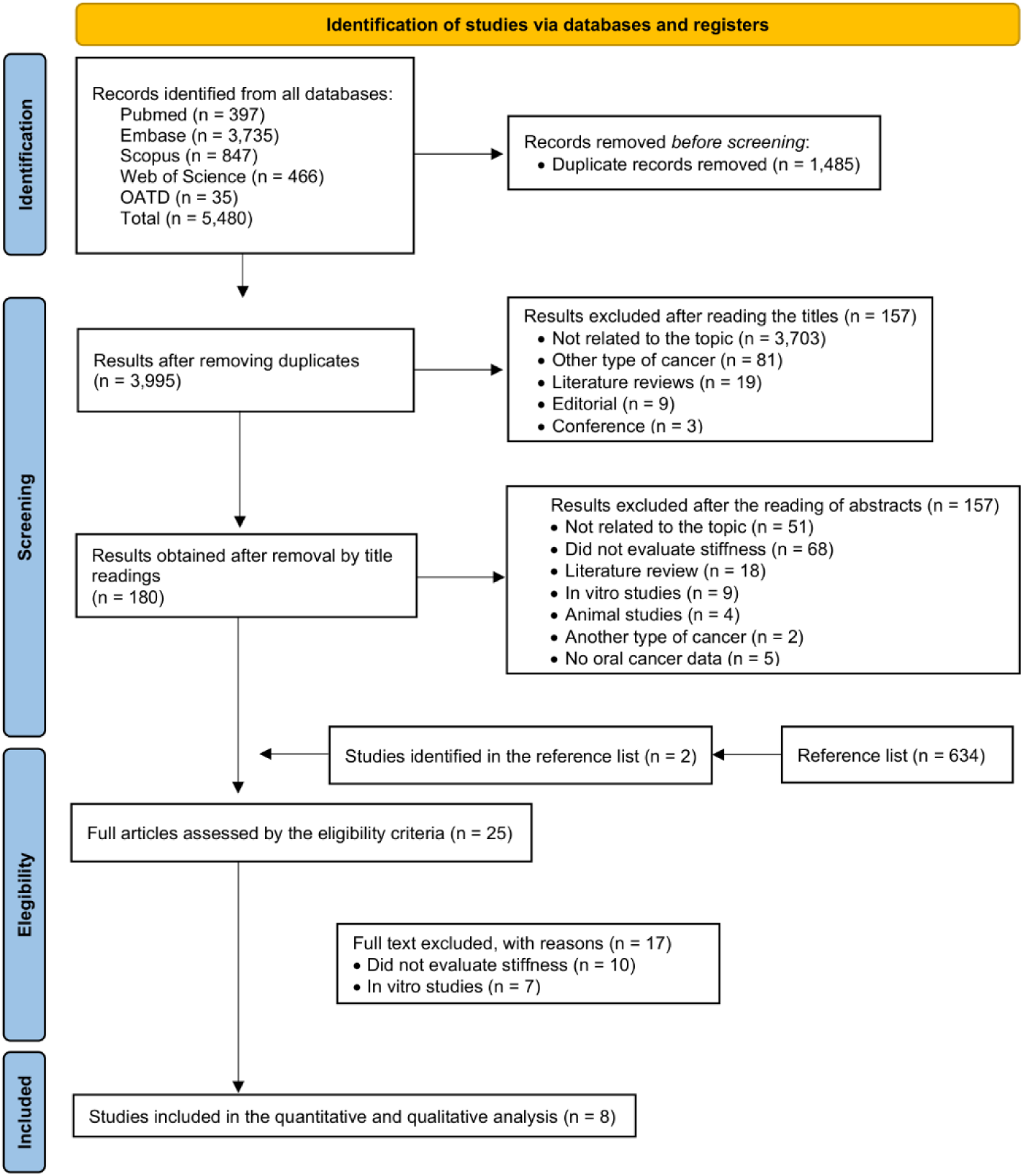
PRISMA flow diagram showing identification, screening, and inclusion of studies in the systematic review and meta-analysis of ECM stiffness in OSCC.

### Study characteristics

The publication dates of the articles ranged from 1995 to 2025, and yielding a total of 436 patients across all studies [27–34] (Table 2), with individual study samples ranging from 2 to 127 patients. The age of the participants ranged from 20 to 91 years, with mean ages varying from 45.2 to 60.2 years when reported. One study [30] additionally included carcinoma *in situ* (CIS), and one study [34] included a mixed sample of malignant and benign oral lesions. The tongue, particularly the lateral border, was the most commonly investigated anatomical location, being the exclusive focus in five studies [28–32]. One study [27] included multiple oral sites (tongue, floor of mouth, and buccal mucosa), and one study did not specify the exact oral location [34].

**Table 2.** Summary of the main characteristics of the eligible studies.

| *Author and year* | *Sample (n)* | *Sex* | *Age (mean)* | *Tumor location* | *Type of tumor* | *Tumor stage* | *Follow-up* | *Stiffness measurement method* | *Stiffness classification* | *Control values* |
| --- | --- | --- | --- | --- | --- | --- | --- | --- | --- | --- |
| *Sato et al., 1995[27]* | 32 | -- | -- | Oral mucosa (Tongue, floor of mouth, buccal mucosa) | OSCC | -- | No | Manual hardness measuring device (2mm probe, 2mm depth, load measurement) | Quantitative (gf) | Normal mucosa |
| *Ogura et al., 1998[28]* | 45 | 31 (male)<br>14 (female) | 26-85 (55.5) | Tongue | OSCC | T1, T2 and T3 | No | Manual hardness measuring device (2mm probe, 2mm depth, strain gauge) | Quantitative (g) | Contralateral normal tongue |
| *Ogura et al., 2000[29]* | 53 | 34 (male)<br>19 (female) | 23-85 (56.1) | Tongue | OSCC | T1, T2, N0 and N1-N2 | 2 years | Manual hardness measuring device (same method) | Quantitative (g) | Contralateral normal tongue |
| *Shingaki et al., 2017[30]* | 17 | 11 (male)<br>6 (female) | 38-85 (--) | Tongue | OSCC and CIS | cT1-T2, N0 | 1 year | Strain elastography (intraoral probe, 7-13 MHz, manual compression, acoustic coupling gel) | Elasticity score 1-4* | Normal tongue (adjacent tissue) |
| *Ogura et al., 2018[31]* | 2 | 2 (male) | 50-59 (--) | Tongue | OSCC | -- | No | Strain elastography (intraoral ultrasonography, Aplio 300, 14 MHz, manual compression) | Quantitative; Strain (%) | -- |
| *Shibata et al., 2021[32]* | 62 | 28 (male)<br>34 (female) | 28-91 (60.2) | Tongue | OSCC | pT1 and pT2 | No | Strain elastography (intraoral, Aplio 500, 7-14 MHz hockey-stick probe, manual compression) | SR and semi-quantitative histology (%CFC, %CCC, %TIICs) | Normal tongue |
| *Jingyuan et al., 2023[33]* | 127 | -- | >60 and <60 (--) | Oral cavity | OSCC | T1-T4; N0-N+ | 3 years | Masson's trichrome staining of collagen fibers in tumor stroma | Semi-quantitative score; Percentage of collagen fibers in the tumor stroma classified into scores | Tumor adjacent epithelium |
| *Babkair, 2025[34]* | 50 | 30 (male)<br>20 (female) | 20-70 (45.2) | Oral lesions (suspicious) | Several | -- | No | Ultrasound elastography (7-15 MHz linear transducer) | SR and Young's Modulus (kPa) | Normal tissue |

-- Not informed by the author; OSCC: oral squamous cell carcinoma; CIS: carcinoma in situ.; \*Grade 1: (very soft; high elasticity): lesion appeared predominantly green to red (70% of total lesion area), with a few small blue areas; Grade 2 (moderately soft; moderately high elasticity): more green areas than blue areas, but not predominant (50–70% of total lesion area); Grade 3 (moderately hard; moderately low elasticity): more blue areas than green areas in the lesion, but not predominant; Grade 4 (very hard; low elasticity): predominantly blue areas in the lesion with a few small green areas; g: gama-force; g:gram; kPa: kilopascal.

Baseline tumor stage was reported in five [28–30,32,33] of the eight studies. Two studies included T1 and T2 tumors, one study [28] also included T3 tumors, one study [32] reported pathological staging (pT1 and pT2), and one study [33] included the full spectrum from T1 to T4. Nodal status was specified in two studies [29,30], with Ogura et al. (2000) [29] reporting N0 to N2 stages and Shingaki et al. (2017) [30] including only N0 patients. Only three studies [29,30,33] reported follow-up periods: Shingaki et al. (2017) [30] followed patients for 1 year, Ogura et al. (2000) [29] for 2 years, and Jingyuan et al. (2023) [33] for 3 years.

Regarding stiffness measurement methods, the most common approach was strain elastography, employed in four studies [30–32,34]. These studies used intraoral ultrasonography with transducer frequencies ranging from 7 to 15 MHz, applying manual compression with acoustic coupling gel or polymer gel to assess tissue elasticity. Three studies [27–29] utilized a manual hardness-measuring device developed by the authors, consisting of a handheld probe (2 mm diameter) pressed into the mucosa to a depth of 2 mm, with load measurement via a strain gauge. One study [33] employed an indirect method, using Masson’s trichrome histological staining to semi-quantify collagen fiber content as a surrogate marker for tissue stiffness.

Stiffness classification varied across studies. Three studies [27–29] used quantitative measurements expressed in grams [28,29] or gamma-force [27]. Three studies [30,32,34] employed elastography-derived metrics, including strain ratio and Young’s modulus. One study [31] expressed results as percentage strain values. One study [33] used a semi-quantitative scoring system (0-4) based on the percentage of collagen fibers visualized in tumor stroma, with a binary classification of low stiffness (score <3) versus high stiffness (score ≥3). The last study [34] explicitly stated the use of arbitrary values to test diagnostic accuracy, which significantly limited the validity of its findings.

### Risk of bias assessment

Based on the proportion of “yes” responses as described in the Methods, only one study [33] was classified as having a low risk of bias, whereas seven studies were classified as having a moderate risk of bias. No study was judged to have a high risk of bias (Table 3). Question 5 of the diagnostic accuracy checklist was considered not applicable for one study [31], as its objective was to characterize and evaluate quantitative strain without defining or applying a diagnostic cutoff to classify lesions.

**Table 3.** Risk of bias assessed by the Joanna Briggs Institute Critical Appraisal Tools for use in JBI Systematic Reviews for Diagnostic Test Accuracy Studies and Cohort.

| *Study design* | *Authors* | *Q.1* | *Q.2* | *Q.3* | *Q.4* | *Q.5* | *Q.6* | *Q.7* | *Q.8* | *Q.9* | *Q.10* | *Q.11* | *% yes/risk* |
| --- | --- | --- | --- | --- | --- | --- | --- | --- | --- | --- | --- | --- | --- |
| **Accuracy** | *Shingaki et al., 2017[30]* | X | √ | X | U | X | √ | U | √ | √ | √ | -- | 50% Moderate |
|  | *Ogura et al., 2018[31]* | X | √ | U | U | NA | √ | U | √ | √ | √ | -- | 50 % Moderate |
|  | *Babkair, 2025[34]* | U | √ | √ | √ | X | √ | U | U | √ | √ | -- | 60% Moderate |
|  | *Ogura et al., 1998[28]* | U | X | √ | U | √ | √ | U | √ | √ | √ | -- | 60% Moderate |
|  | *Ogura et al., 2000[29]* | U | √ | √ | U | X | √ | U | √ | √ | √ | -- | 60% Moderate |
|  | *Shibata et al., 2021[32]* | X | √ | √ | U | X | √ | U | √ | √ | √ | -- | 60% Moderate |
| **Cohort** | *Sato et al., 1995[27]* | U | X | √ | U | X | √ | √ | √ | √ | √ | -- | 60% Moderate |
|  | *Jingyuan et al., 2023[33]* | √ | √ | √ | √ | √ | √ | √ | U | U | U | √ | 72% Low |

**Diagnostic Test Accuracy Studies** - Q1. Was a consecutive or random sample of patients enrolled?; Q2. Was a case control design avoided?; Q3 Did the study avoid inappropriate exclusions?; Q4 Were the index test results interpreted without knowledge of the results of the reference standard?; Q5 If a threshold was used, was it pre-specified?; Q6 Is the reference standard likely to correctly classify the target condition?; Q7 Were the reference standard results interpreted without knowledge of the results of the index test?; Q8 Was there an appropriate interval between index test and reference standard?; Q9 Did all patients receive the same reference standard?; Q10 Were all patients included in the analysis?
**Cohort Studies** - Q.1 Were the two groups similar and recruited from the same population?; Q.2 Were the exposures measured similarly to assign people to both exposed and unexposed groups?; Q.3 Was the exposure measured in a valid and reliable way?; Q.4 Were confounding factors identified?; Q.5 Were strategies to deal with confounding factors stated?; Q.6 Were the groups/participants free of the outcome at the start of the study (or at the moment of exposure)?; Q.7 Were the outcomes measured in a valid and reliable way?; Q.8 Was the follow up time reported and sufficient to be long enough for outcomes to occur?; Q.9 Was follow up complete, and if not, were the reasons to loss to follow up described and explored?; Q.10 Were strategies to address incomplete follow up utilized?; Q.11 Was appropriate statistical analysis used?
√ - yes; X - No; U – Unclear; NA – Not Applicable.

### Individual study results

Of the eight studies [27–34], only three [28,30,32] reported mean tumor thickness, which ranged from 3.0 to 4.4 mm (Table 4). Stiffness parameters were assessed using diverse units and methodologies across studies, including gram-force (gf) [27], grams (g) [28, 29], kilopascals (kPa) [34], percentage strain [31], and semi-quantitative scores [30, 33]. One study [31] reported a complete absence of strain (0%) in tumors, whereas another classified stiffness based on collagen content, with >50% collagen fibers indicating high stiffness [33].

**Table 4.**
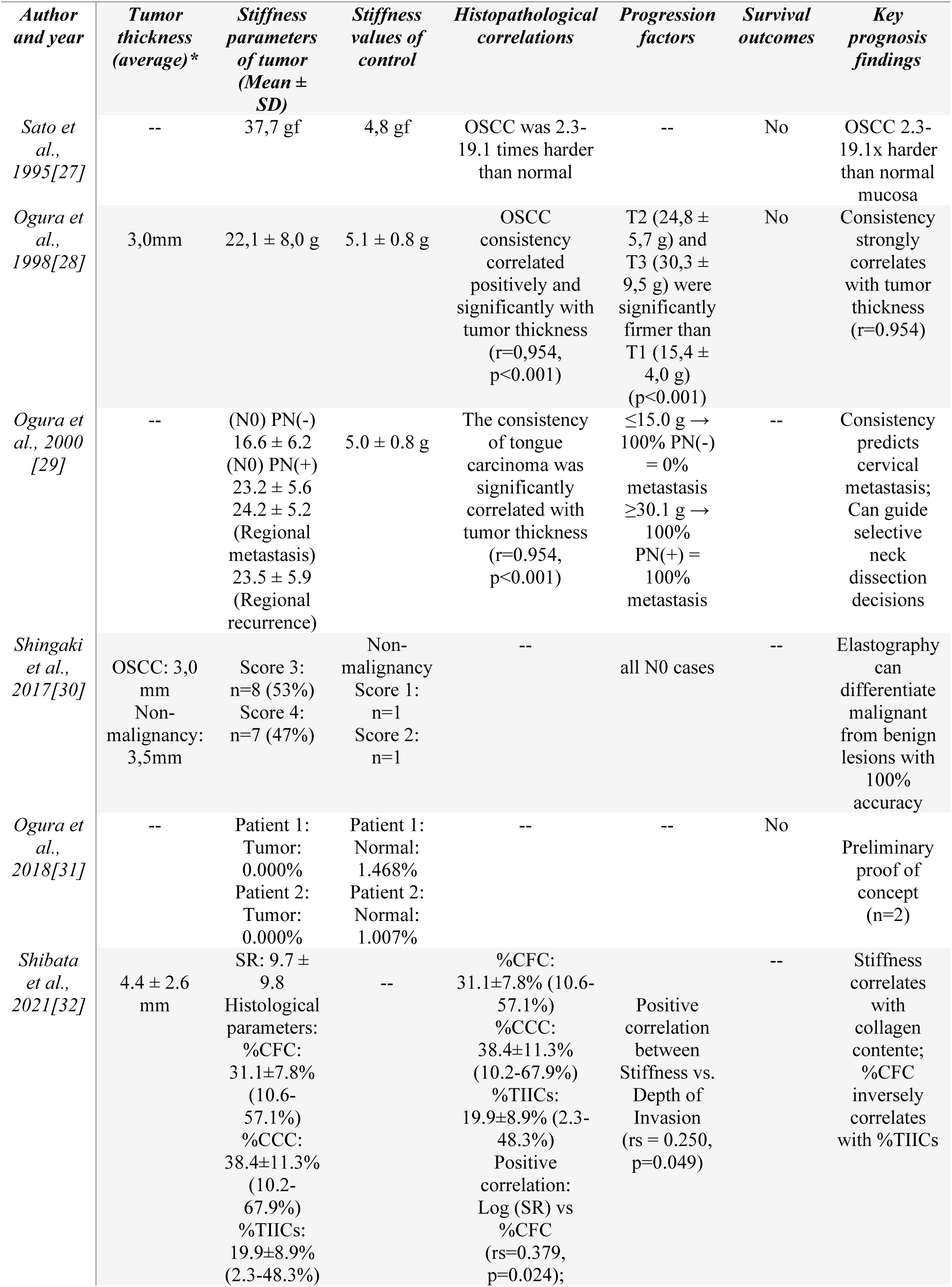
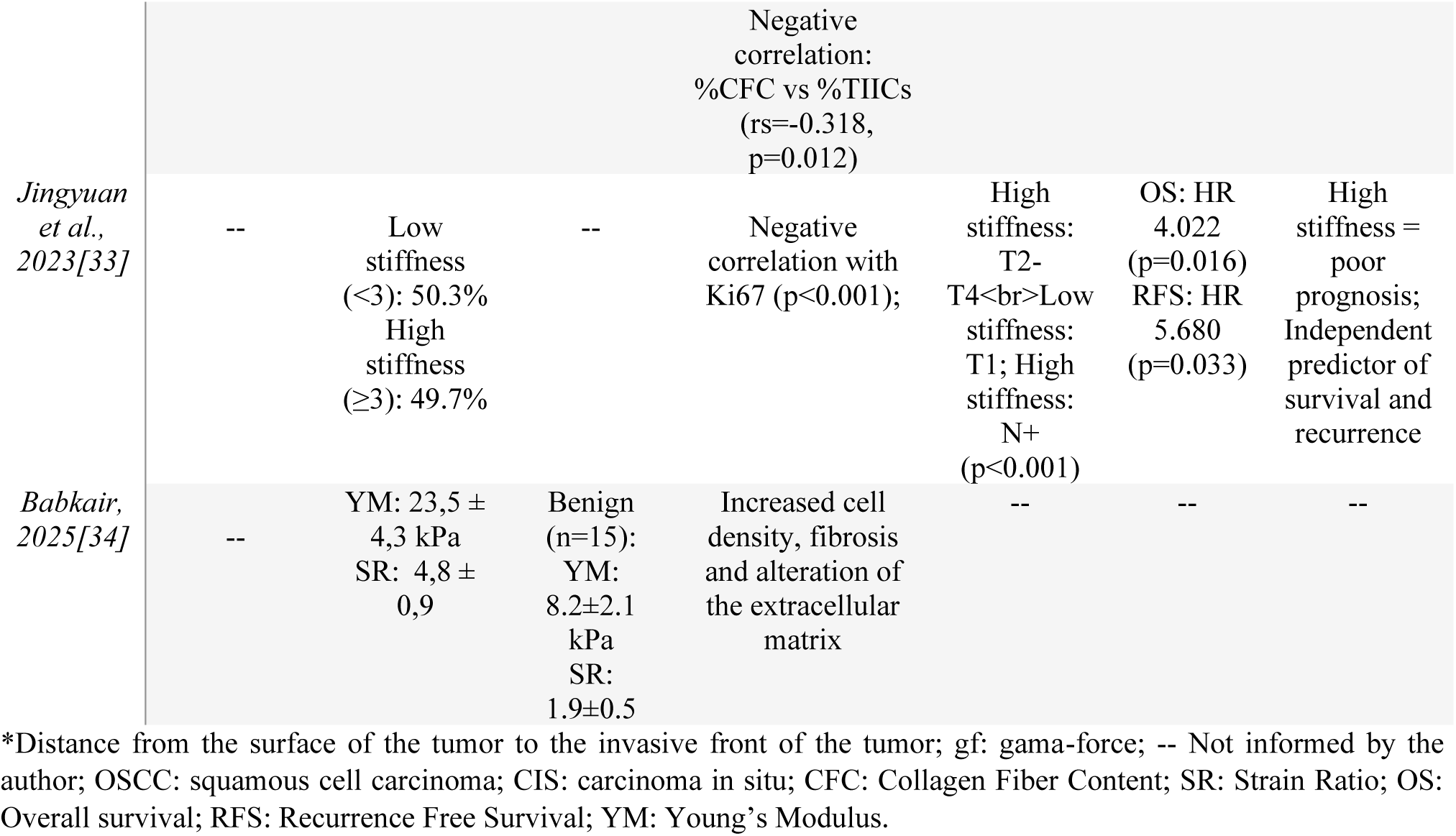
Summary of the main results of the eligible studies.

One study [27] reported a mean stiffness of 37.7 gf for OSCC, which was 2.3 to 19.1 times higher than that of normal mucosa (4.8 gf), with a significant difference (p < 0.001). Another study [28] found an overall mean consistency of 22.1 ± 8.0 g, with values varying significantly according to T stage: T1 tumors measured 15.4 ± 4.0 g (n = 17), T2 tumors 24.8 ± 5.7 g (n = 21), and T3 tumors 30.3 ± 9.5 g (n = 7), with T2 and T3 tumors being significantly firmer than T1 tumors (p < 0.001). In a subsequent study [29], tumors with pathologically negative nodes (PN−) measured 16.8 ± 6.3 g (n = 36), whereas those with regional metastasis measured 24.2 ± 5.2 g (n = 7) and those with regional recurrence measured 23.5 ± 5.9 g (n = 10); both metastatic and recurrent tumors were significantly firmer than PN− tumors (p < 0.05). Conversely, some studies employed strain elastography to report strain ratios (SR) [32,34], Young’s modulus values [34], or percentage strain [31]. Mean strain ratios ranged from 4.8±0.9 to 9.7±9.8 for OSCC across studies [32,34], compared to benign lesions with values of 1.9±0.534. Young’s modulus for malignant lesions was 23.5±4.3 kPa compared to 8.2±2.1 kPa for benign lesions (p<0.001) [34]. One preliminary study [31] reported percentage strain values demonstrating complete absence of strain (0.000%) in tumor tissue.

Finally, qualitative and semi-quantitative scoring systems were employed in two studies [30,33]. One study [30] utilized a four-grade elasticity scoring system based on color-coded elastography images, where scores ranged from 1 (very soft, predominantly green-red appearance) to 4 (very hard, predominantly blue appearance). In this study, all carcinomas (n=15) scored 3 (n=8, 53%) or 4 (n=7, 47%), while all non-malignant lesions (n=2) scored 1 or 2, achieving perfect diagnostic separation (100% sensitivity and specificity). Another study [33] employed Masson’s trichrome staining to semi-quantify collagen fiber content in tumor stroma as an indirect measure of matrix stiffness, using a scoring system from 0 to 4 based on the percentage of collagen fibers: 0 (<10%), 1 (11-30%), 2 (31-50%), 3 (51-80%), and 4 (>80%).

Beyond stiffness measurements, six studies [28–30,32–34] investigated histopathological correlations in relation to tumor stiffness, of which three [28, 29, 32] demonstrated significant positive correlations between OSCC stiffness and tumor thickness. Only one study [33] reported long-term survival outcomes. In that study, patients with high matrix stiffness (Masson score ≥3) had significantly shorter overall survival (OS) and recurrence-free survival (RFS) compared with those with low stiffness (score <3) (log-rank test, p < 0.001 for OS and p = 0.047 for RFS). Multivariate Cox regression analysis, adjusting for other clinical and pathological variables, identified high matrix stiffness as an independent risk factor for OS (hazard ratio [HR] = 4.022; 95% CI: 1.294–12.495; p = 0.016) and RFS (HR = 5.680; 95% CI: 1.146–28.143; p = 0.033). The remaining seven studies [27–32,34] did not assess or report survival outcomes.

### Meta-analysis results

A meta-analysis of five studies comprising 307 patients was conducted to assess the standardized mean difference in tissue stiffness between OSCC and normal oral mucosa. The included studies were published between 1995 and 2025, with individual study samples ranging from 32 to 127 participants. The pooled analysis revealed a statistically significant increase in stiffness in OSCC tissue compared with normal oral mucosa, with a standardized mean difference (Cohen’s d) of 5.28 (95% CI: 2.61–7.94; Z = 3.88; p < 0.001) (Figure 2A). All individual study estimates favored higher stiffness in OSCC, with confidence intervals that did not cross zero.

**Figure 2.**
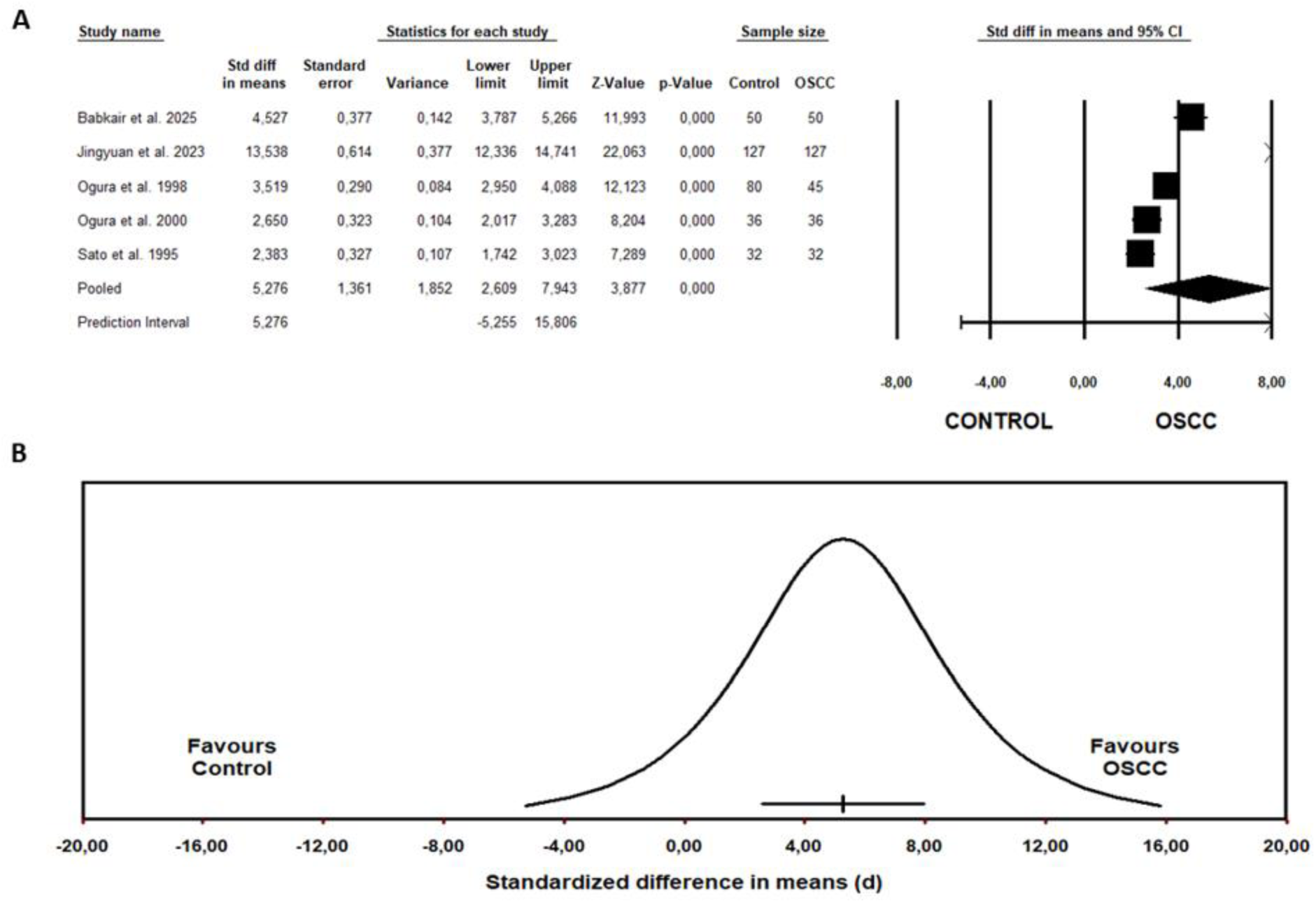
Forest plot and distribution of true effects for extracellular matrix stiffness in oral squamous cell carcinoma versus normal oral mucosa; **(A)** Forest plot showing standardized mean differences (Cohen’s d) with 95% confidence intervals for five quantitative studies comparing extracellular matrix stiffness between oral squamous cell carcinoma and normal oral control samples. Individual study estimates are represented by squares, with horizontal lines indicating 95% confidence intervals. The pooled effect estimate (random-effects model, DerSimonian–Laird method) is shown as a diamond, indicating a standardized mean difference of 5.28 (95% CI: 2.61–7.94; p < 0.001). The prediction interval (−5.26 to 15.81) reflects the range of true effects expected in future studies. The vertical reference line at zero indicates no difference between groups. All individual studies and the pooled estimate favor higher stiffness in oral squamous cell carcinoma (right side of zero); **(B)** Distribution plot of true effects illustrating the probability density of standardized mean differences across studies. The bell-shaped curve represents the distribution of true effect sizes, with the majority of effects concentrated in the positive range (favoring oral squamous cell carcinoma), indicating that extracellular matrix stiffness is elevated in oral squamous cell carcinoma across studies, with a uniform direction of effect. The x-axis ranges from −20.00 to 20.00 standardized difference units, with directional indicators showing “Favors Control” (left) and “Favors OSCC” (right). The distribution illustrates the substantial heterogeneity among studies (I² = 98.6%), with a uniform direction of effect favoring higher stiffness in oral squamous cell carcinoma.

Substantial statistical heterogeneity was observed, with an I² statistic of 98.6%, indicating that most of the variability in effect sizes was due to genuine differences between studies rather than chance. This heterogeneity was anticipated and likely reflects the diversity in stiffness measurement methodologies (including elastography, manual devices, and histological quantification), as well as variations in study design, patient populations, tumor characteristics (including tumor thickness ranging from 3.0 to 4.4 mm), and anatomical locations (predominantly tongue). Given this heterogeneity, a random-effects model (DerSimonian–Laird method) was used. The 95% prediction interval for the true effect ranged from −5.25 to 15.81, reflecting the considerable variability in effect sizes across studies while still suggesting that future studies are likely to observe higher stiffness in OSCC tissue on average (Figure 2B).

Potential publication bias was assessed using funnel plots, precision plots, Egger’s regression intercept test, and Begg’s rank correlation test (Figure 3). Funnel plot and precision plot analyses showed a roughly symmetric distribution of studies around the pooled effect estimate. Egger’s and Begg’s tests did not detect statistically significant asymmetry. However, given the small number of studies included in the meta-analysis, these tests have limited power, and the presence of publication bias cannot be ruled out.

**Figure 3.**
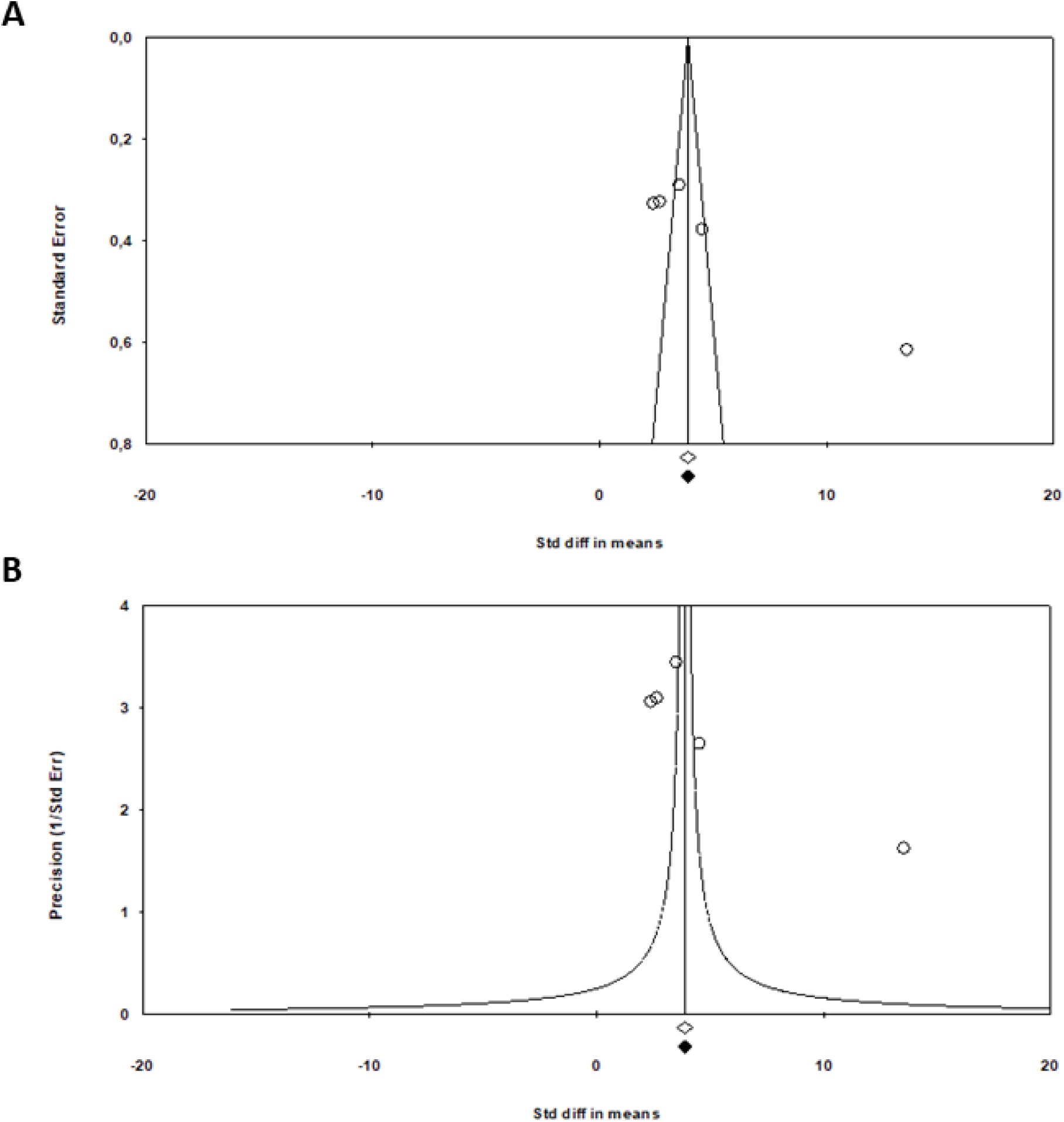
Funnel plot and precision plot for assessment of publication bias; **(A)** Funnel plot displaying the relationship between standardized mean differences (x-axis: “Std diff in means,” ranging from −20 to 20) and standard error (y-axis, ranging from 0 to ∼3.5). Each circle represents an individual study, with the central diamond indicating the pooled effect estimate (Cohen’s d = 5.28). The funnel-shaped contour lines represent the expected 95% confidence region for studies assuming no publication bias. The distribution of studies around the pooled effect did not reveal obvious asymmetry suggestive of publication bias. Studies are distributed across various levels of precision, with larger studies (lower standard error) positioned toward the apex of the funnel and smaller studies (higher standard error) distributed more broadly at the base; **(B)** Precision plot presenting the inverse relationship between precision (y-axis: “Precision [1/Std Err],” ranging from 0 to ∼10) and standardized mean differences (x-axis: “Std diff in means,” ranging from −20 to 20). Each circle represents an individual study, with the central diamond indicating the pooled effect estimate. The funnel-shaped contour lines define the expected symmetric distribution region under the assumption of no publication bias. The symmetric arrangement of data points around the pooled effect does not suggest preferential publication of studies with larger or smaller effect sizes. Higher precision values (upper portion) represent larger studies with smaller standard errors, while lower precision values represent smaller studies with larger standard errors. Quantitative tests (Egger’s regression intercept test and Begg’s rank correlation test) did not detect statistically significant funnel plot asymmetry (p > 0.05), although the power to detect publication bias is limited with only five studies.

## DISCUSSION

Increased tissue stiffness is a characteristic property of tissues and organs undergoing morphological and functional alterations [35]. Recent studies have evaluated tumor stiffness as a diagnostic tool and as a method for predicting metastatic potential. This systematic review synthesized evidence from eight studies involving patients with OSCC, indicating that ECM stiffness is elevated in tumor tissue compared with normal oral mucosa and may represent a potentially clinically relevant biomechanical marker of malignant transformation in the oral cavity. The meta-analysis showed that tumor tissue exhibits higher stiffness compared with normal oral mucosa, and one study reported that high matrix stiffness was associated with worse overall survival and recurrence-free survival. Here we discuss the primary reliance on elastography for stiffness measurements versus other methods, the quantitative stiffness values for OSCC and other tissues, stiffness-mediated signaling, and measurement limitations.

OSCC is a tumor of malignant epithelial origin that, in most cases, presents clinically as a hardened solid mass [9], which is closely related to qualitative and quantitative alterations in ECM components [9,10]. Among the most evaluated methods to measure such stiffness, elastography was employed in four studies [30–32,34] of the present systematic review. This is an approach that visualizes and quantifies tissue elasticity through ultrasound evaluation of differential displacement under stress or by shear waves and mechanical waves [10]. This approach enables the measurement of quantitative metrics (strain ratio, Young’s modulus) that facilitate objective comparison between patients and tissues and has been previously used across breast tumors [9], liver [36,37], pancreas [38,39], and prostate cancers [40,41]. Supporting the validity of this measurement method, Shingaki et al.[30] demonstrated difference between malignant (elasticity scores 3–4) and benign (scores 1–2) oral lesions, while Shibata et al.[32] correlated strain ratios with histopathological collagen content, thereby validating the biological basis of elastography measurements. However, it is important to acknowledge that elastography requires specialized equipment, technical expertise, and careful attention to compression uniformity to ensure reproducible and reliable results [9].

Nevertheless, the heterogeneity of stiffness measurement approaches employed in the included studies, such as manual mechanical devices [27–29], strain elastography [30–32,34], and histological quantification of collagen content [33], reflects the evolving landscape of biomechanical tissue characterization for tumor analyses. Manual hardness-measuring devices, evaluated in three studies [27–29], provided objective quantification of tissue resistance to deformation through controlled probe displacement and load measurement. Although operator-dependent and requiring direct tissue contact, these devices demonstrated good reproducibility [27–29]. The simplicity and low cost of these devices may make them particularly suitable for resource-limited settings or for quantifying intraoperative manual palpation [27–29]. On the other hand, histological quantification of collagen fibers through Masson’s trichrome staining, as employed in one study [33], provides an indirect, proxy assessment of matrix stiffness [33]. The semi-quantitative nature of this histological scoring may not capture continuous biological variation, and post-fixation processing may alter tissue mechanical properties, limiting direct inference for in vivo stiffness [42]. Nevertheless, these approaches are accessible to researchers without the specialized equipment required for elastography measurements.

The quantitative stiffness values for OSCC in this systematic review ranged approximately from 15–40 g in manual measurements, 4.8–9.7 in strain ratios, and 23.5 kPa in Young’s modulus. In comparison, studies of breast carcinomas have reported mean Young’s modulus values ranging from 80–120 kPa measured by shear wave elastography [43], and a recent study reported mean hardness values of 189.5 kPa for breast tumors, higher than those observed in OSCC. This difference may be attributed to the composition and organization of the ECM in breast carcinomas, which exhibit extensive desmoplasia with considerable deposition of type I collagen organized in parallel linear bundles [45]. In contrast, although OSCC demonstrates significant collagen deposition, the fibrillar architecture tends to be more disorganized and multidirectional, particularly at invasive margins [46]. Studies [47] have demonstrated mean hardness exceeding 100 kPa in pancreatic tumors, also significantly higher than that found in OSCC in the present review. Similarly, liver tumors, specifically hepatocellular carcinoma [48], demonstrated stiffness of approximately 55 kPa, while studies [49] evaluating prostate tumors reported values of 91.6–131.8 kPa [48]. For other tumors such as cervical squamous cell carcinomas (CSCC), which share histological and etiological characteristics with OSCC, mean stiffness has been reported as 5.27 kPa when evaluated with magnetic resonance elastography [53]. Additionally, esophageal squamous cell carcinomas have demonstrated strain ratios by ultrasound elastography of 1 ± 0.9 (range 0.3–2.9) [54], which are numerically lower than, but broadly comparable in direction to, the strain ratios reported for tongue cancer in this review. Multiple factors are likely involved in these differences, including variable stroma–tumor proportion [50], specific ECM composition of each tumor [51,52], and methodological heterogeneity in stiffness measurement.

When considering the relationship between stiffness and the degree of tumor invasion across sites, pancreatic and breast carcinomas, which frequently exhibit early systemic dissemination, demonstrate very high absolute stiffness [56, 57]. In contrast, basal cell carcinomas of the skin, known for their high local invasion capacity but low metastatic potential, show moderately elevated stiffness compared with normal dermis [58]. OSCC occupies an intermediate position, with significant locoregional invasiveness and cervical lymph node metastasis, but less frequent distant dissemination than breast or pancreatic cancers [59]. The finding from one study [29] in this systematic review that tumors exceeding 30.1 g showed 100% lymph node involvement is consistent with the hypothesis that higher stiffness correlates with invasive capacity, potentially mediated through mechanotransduction pathways that promote epithelial-mesenchymal transition, matrix metalloproteinase expression, and migratory phenotypes [60].

Understanding how physical properties of the ECM are transformed into biological responses is fundamental to comprehending the mechanisms that influence tumor progression [60]. The focal adhesion kinase (FAK) pathway and Hippo-YAP/TAZ signaling are currently considered the main transducers of mechanical signals, with matrix stiffness promoting FAK phosphorylation at focal adhesions and nuclear translocation of YAP/TAZ, driving expression of pro-proliferative and pro-invasive genes. Based on this, one study [33] from the present systematic review demonstrated that mechanical stress related to stiffness causes nuclear envelope rupture and DNA damage, which is sensed by cyclic GMP-AMP synthase (cGAS), triggering sequential phosphorylation of STING (stimulator of interferon genes), TBK1, and IRF3, creating dormant cell populations and favoring epithelial-mesenchymal transition, fundamental for tumor progression.

Despite limitations in the included studies, such as heterogeneity in stiffness measurement methods and differences in protocols, the findings of this systematic review may have relevant clinical implications. The substantial heterogeneity observed in this meta-analysis (I² = 98.6%) reflects the methodological diversity of stiffness measurement approaches employed in the included studies, including manual mechanical devices [27–29], strain elastography [30–32,34], and histological quantification of collagen content [33]. While this heterogeneity indicates that the direction of the association (higher stiffness in OSCC than in normal mucosa) is consistent across diverse measurement techniques, it limits direct comparison of absolute stiffness values. Standardization of measurement protocols and reporting formats would facilitate future meta-analyses and clinical translation. Variations in tumor characteristics (thickness: 3.0–4.4 mm) and anatomical locations (predominantly tongue) may also contribute to effect size heterogeneity. The tongue, particularly the lateral border, represents the most common OSCC site and may exhibit distinct biomechanical properties compared to other oral sites. Future research should systematically investigate these variables to identify potential moderators of effect size.

Overall, this systematic review and meta-analysis provide consistent evidence that ECM stiffness is significantly elevated in OSCC compared with normal oral mucosa (standardized mean difference = 5.28; 95% CI: 2.61–7.94; p < 0.001). The consistency of this finding across diverse measurement methodologies and study populations, together with the absence of detectable funnel plot asymmetry (acknowledging the limited power of these tests), supports the biological relevance of ECM stiffness as a characteristic feature of oral malignancy. The universal elevation of tumor tissue stiffness across all included studies, coupled with documented correlations between ECM stiffness and clinically relevant parameters (tumor thickness, lymph node metastasis, and, in one study, survival outcomes), supports ECM stiffness as a potentially relevant biomechanical marker in OSCC.

The integration of biomechanical assessment into clinical practice could potentially enhance diagnostic accuracy, improve prognostic stratification, and facilitate more personalized therapeutic guidance. However, clinical translation will require standardization of measurement protocols, prospective validation studies with adequate sample sizes, and systematic investigation of the mechanistic relationships linking ECM stiffness to tumor progression and clinical outcomes. At present, the use of ECM stiffness assessment in OSCC remains more promising in research settings than in routine clinical practice. Ultimately, the integration of ECM stiffness assessment into comprehensive diagnostic and prognostic frameworks could contribute to earlier detection, improved risk stratification, and more effective and personalized management of patients with OSCC.

## CONCLUSION

Tumor tissue in OSCC exhibited higher stiffness than normal oral mucosa in all included studies, with associations reported between higher stiffness, tumor thickness, lymph node metastasis and, in one study, worse survival. These findings support ECM stiffness as a promising biomechanical marker in OSCC, although further standardized and prospective studies are needed before clinical implementation.

## Approval committee/the internal review board (IRB)

not applicable

## Clinical trial number

not applicable.

## Human Ethics and Consent to Participate declarations

not applicable

## Consent to Participate

not applicable

## Data Availability

All data produced in this study are available upon reasonable request to the authors

## FUNDING DECLARATION

The research leading to these results received funding from Conselho Nacional de Desenvolvimento Científico e Tecnológico, Brazil (406258-2022-8 to M.L.L) and the U.S. National Institutes of Health (R01CA206880 to A.J.E. and R01CA280279 to A.J.E. and M.L.L.

## CONFLICT OF INTEREST

The authors declare that they have no known competing financial or personal interests that could have appeared to influence the work reported in this paper.

## ACKNOWLEDGEMENTS

The authors acknowledge funding from Conselho Nacional de Desenvolvimento Científico e Tecnológico (406258-2022-8 to M.L.L) and the U.S. National Institutes of Health (R01CA206880 to A.J.E. and R01CA280279 to A.J.E. and M.L.L.) and a cy pres research award from the Krueger v. Wyeth settlement fund (to A.J.E.).

## AUTHORS’ CONTRIBUTIONS

Igor Felipe Pereira Lima: Conceptualization, Methodology, Investigation, Data curation, Formal analysis, Software, Writing – original draft, Writing – review & editing.

Ana Carolina Toebe Silva: Investigation, Methodology, Writing – original draft.

Adam Jeffrey Engler: Funding acquisition, Investigation, Visualization, Writing – review & editing.

Leonardo da Silva Bittencourt: Data curation, Formal analysis, Methodology, Software, Writing – review & editing.

Marcelo Lazzaron Lamers: Conceptualization, Investigation, Supervision, Project administration, Funding acquisition, Writing – review & editing.

All authors contributed to the interpretation of the findings, critically revised the manuscript, and approved the final version.

## Notes

### Competing Interest Statement

The authors have declared no competing interest.

